# Repurposing of rituximab biosimilars to treat B cell mediated autoimmune diseases

**DOI:** 10.1101/2023.09.22.23295633

**Authors:** Agata Mostkowska, Guy Rousseau, Noël J-M Raynal

## Abstract

Rituximab, the first monoclonal antibody approved for the treatment of lymphoma, eventually became one of the most popular and versatile drugs ever in terms of clinical application and revenue. Since its patent expiration, and consequently, the loss of exclusivity of the original biologic, its repurposing as an off-label drug has increased dramatically, propelled by the development and commercialization of its many biosimilars. Currently, rituximab is prescribed worldwide to treat a vast range of autoimmune diseases mediated by B cells. Here, we present a comprehensive overview of rituximab repurposing in 116 autoimmune diseases across 17 medical specialties, sourced from over 1,530 publications. Our work highlights the extent of its off-label use and clinical benefits, underlining the success of rituximab repurposing for both common and orphan immune-related diseases. We discuss the scientific mechanism associated with its clinical efficacy and provide additional indications for which rituximab could be investigated. Our study presents rituximab as a flagship example of drug repurposing owing to its central role in targeting CD20^+^ B cells in over 100 autoimmune diseases.

## Introduction

The Food and Drug Administration (FDA) approved rituximab for marketing in the United States (US) in 1997 under the brand name RITUXAN. This was a watershed moment in the history of monoclonal antibody (mAb) therapeutics, as rituximab was the first to make its way onto the cancer biologics market.^1^ Although classified as an anticancer agent, rituximab is not considered a chemotherapy but rather an immunotherapy. Developed by IDEC Pharmaceuticals and later acquired by Biogen (which collaborated with Genentech and Hoffman-LaRoche shortly thereafter), rituximab was approved as a single agent for the treatment of relapsed or refractory CD20^+^ B cell follicular lymphoma—a subtype of non-Hodgkin lymphoma.^2^ It was later approved for use in combination with chemotherapeutic agents to sensitize chemoresistant tumor cells to cytotoxic drugs, which improves therapeutic outcomes.^3^

Rituximab is a chimeric mAb engineered from human immunoglobulin (Ig) G constant domains and murine anti-CD20 variable regions. It acts by targeting the cluster of differentiation 20 (CD20) protein, a cell surface antigen specific to B cells. CD20 is expressed in most but not all stages of B cell development. A lack thereof renders hematopoietic stem cells and plasma cells unaffected by rituximab treatment (FIG. 1). The efficacy of rituximab in eliminating CD20^+^ B cells is attributed to its unique three-fold mechanism of action: 1) complement-dependent cytotoxicity, 2) antibody-dependent cell-mediated cytotoxicity, and 3) induction of apoptosis. Rituximab is a type I CD20 mAb with approximately double the CD20 binding capacity than its type II counterpart, and is thought to bind in a way that does not obstruct further antibody binding.^4–8^ Its specificity at inducing B cell death makes rituximab an effective treatment for B cell malignancies.^9^ The central role of B cells in autoimmunity is the common denominator of a wide range of indications and the rationale behind expanding rituximab use across.^2,10–12^

**Fig. 1:**
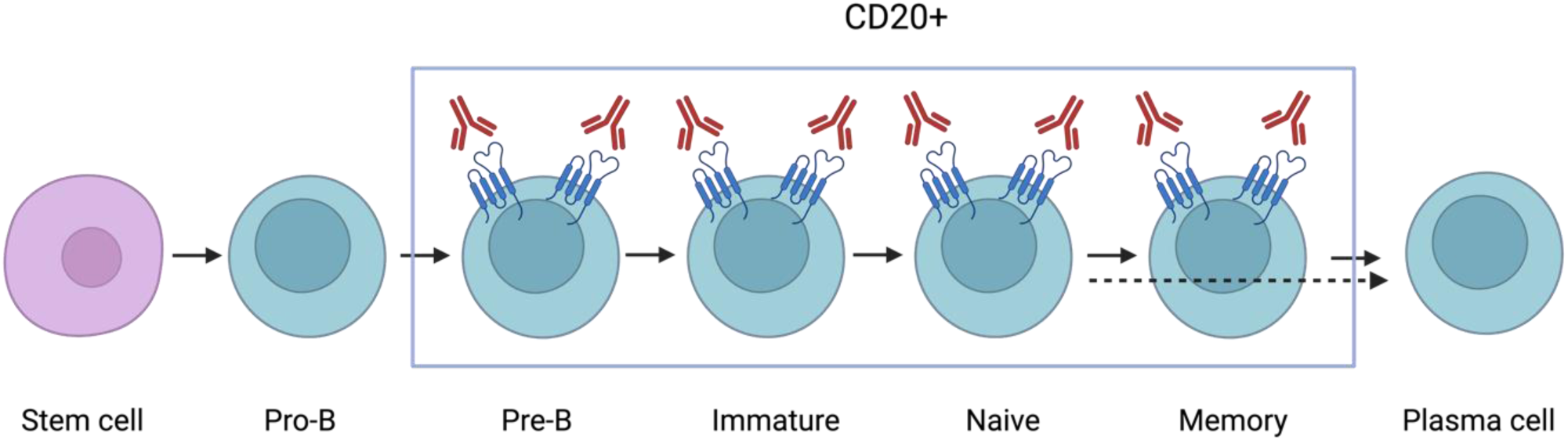
Rituximab targets B cell subtypes that express CD20. B cells first differentiate from bone marrow-derived hematopoietic stem cells into pro-B cells, then develop into pre-B cells, at which point they begin expressing CD20. Pre-B cells differentiate into CD20^+^ immature B cells before leaving the bone marrow and entering the circulation and lymphoid tissue, where they become CD20 naïve B cells. Upon antigen activation, naïve B cells can either differentiate into CD20^+^ memory B cells, then into CD20^-^ plasma cells, or directly differentiate into CD20^-^ plasma cells before migrating back to the bone marrow.

Rituximab’s approved applications gradually expanded to include several blood cancers and autoimmune diseases, namely, chronic lymphocytic leukemia, rheumatoid arthritis (RA), granulomatosis with polyangiitis, microscopic polyangiitis, as well as pemphigus vulgaris (PV). In 1998, it was approved for the same indications by the European Medicines Agency (EMA), under the trade name MabThera. Two years later, Health Canada granted its approval for the same indications (except PV) under the brand name RITUXAN. The increase in approved indications came with a significant rise in global sales.^13^ What started off as the first mAb approved for the treatment of blood and bone-marrow cancer ended up being one of the biggest blockbuster drugs of all time. Rituximab sales increased steadily from 5.5 million USD in 1997 to close to 7.5 billion USD in 2015. Global sales decreased after patents expired in Europe, the US, and Canada in 2013, 2018, and 2019, respectively, with the introduction of biosimilars.^13^ Rituximab’s high demand has ranked it among the top-selling biologics in the world for over 20 years, providing a lucrative market for rituximab biosimilars after patent expiration.^13^ Rituximab use is expected to increase further as biosimilars reduce the price of treatment.^14^ While much of its clinical use is attributed to cancer (64%) and RA (11%) treatment, approximately 25% of total sales stem from off-label prescribing.^13–16^

The use of existing marketed drugs beyond their clinical indication is known as “drug repurposing” or “repositioning.”^17^ There is great interest in repurposing approved medications in lieu of testing novel molecular compounds, as approved drugs have already successfully passed costly toxicological studies, and their pharmacokinetic and pharmacodynamic properties are well characterized. As such, repurposing de-risked drugs for diseases of similar therapeutic molecular targets encourages off-label use. The rationale behind prescribing rituximab biosimilars as off-label treatments for autoimmune and immune-mediated diseases is to abolish self-destructive inflammatory signals mediated by CD20^+^ B cells. In fact, the current literature supports the use of B cell-directed and CD20-targeted therapies, including rituximab, to treat autoimmune diseases.^10,12,18–23^ From a biological standpoint, B cells contribute to autoimmunity in several ways: they not only produce autoantibodies but also act as antigen-presenting cells (APCs), reciprocally interact with T cells, and secrete inflammatory cytokines (FIG. 2).^24^ Fundamentally, the underlying strategy when treating autoimmunity is to restore a normal immune response by eliminating the autoreactive lymphocytes responsible for disease maintenance, progression, and symptom exacerbation.^25^

**Fig. 2:**
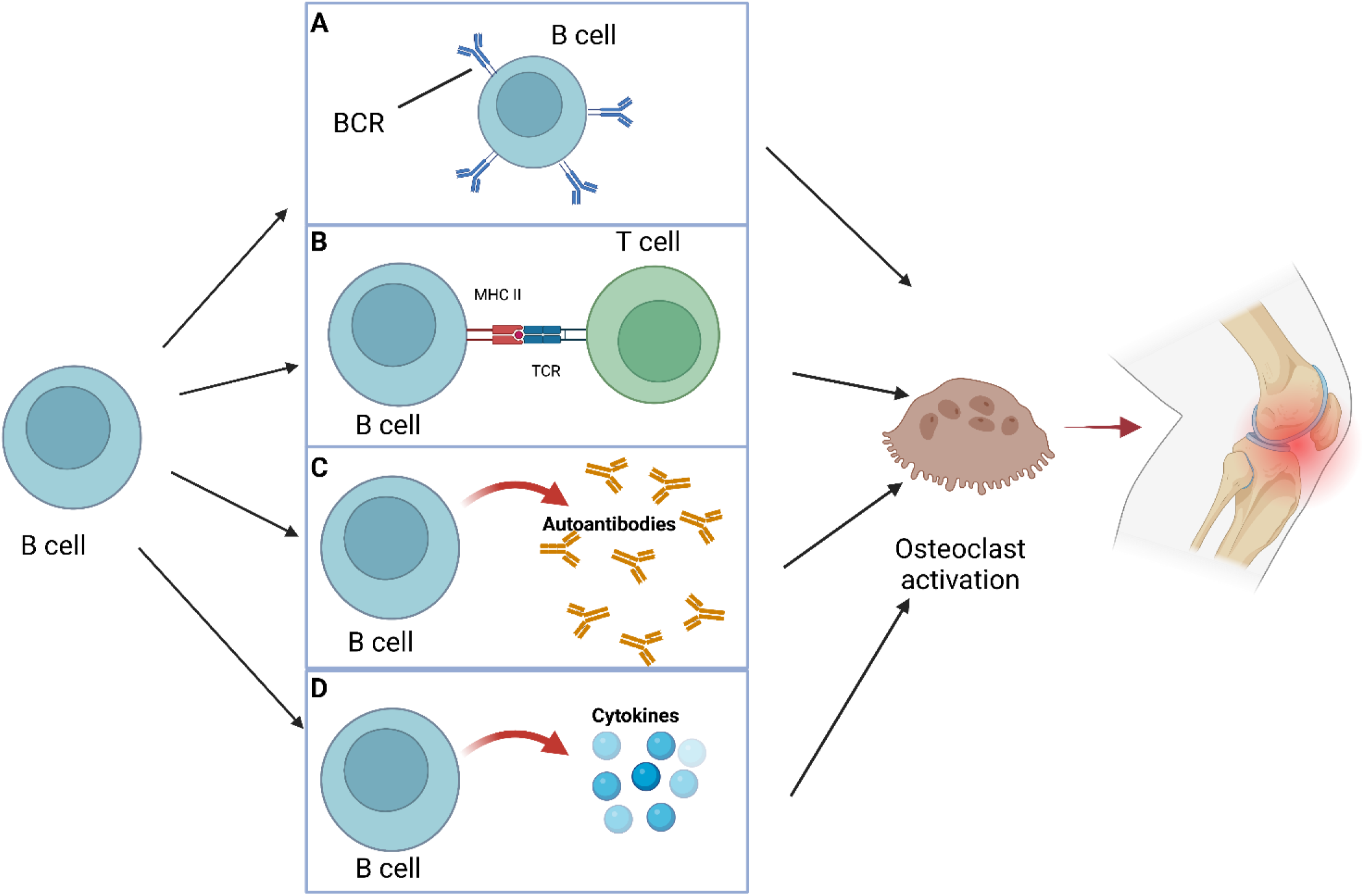
Roles of B cells in (auto)immunity, focusing on RA. Different mechanisms in which B cells are involved and contribute to autoimmune or immune-mediated disorders. B cells perform the following functions: they **A.** act as antigen-presenting cells (APCs) through their B cell antigen receptors (BCRs), **B.** reciprocally interact with T cells through the major histocompatibility complex class II (MHC II) and T cell receptors (TCRs), **C.** produce autoantibodies, and **D.** produce inflammatory cytokines. In RA, these B cell functions lead to osteoclast activation, which perpetuates joint inflammation.

Revolutionary progress has been made in autoimmune disease management through the use of nonsteroidal anti-inflammatory drugs, glucocorticoids, conventional disease-modifying antirheumatic drugs, and newer biologics. However, most autoimmune diseases remain incurable, and providing symptomatic relief and preventing chronic complications caused by inflammatory responses is the current standard of care.^26^ However, many patients are refractory or intolerant to conventional treatments and require alternative therapies. Therapeutic mAbs that target select pathways or components of the immune system may offer more favorable toxicological profiles than non-selective small molecules.^27^ Therefore, immunomodulatory drugs targeting CD20 with demonstrated efficacy and safety profiles have a promising future.^28^ Although rituximab is generally well tolerated, B cell depletion causes side effects predominantly owing to a dampened immune response.^8,29,30^

The purpose of this review is to provide a comprehensive overview of the off-label clinical use of rituximab as a single agent to treat autoimmune and immune-mediated diseases. Our work is structured around the aforementioned roles of B cells in autoimmune diseases across multiple medical specialties, including rheumatology, neurology, dermatology, endocrinology, hematology, ophthalmology, gastroenterology, nephrology, immunology, cardiology, psychiatry, gynecology, pulmonology, hepatology, otolaryngology, urology, and infectiology. In addition, we describe how B cell depletion rationalizes rituximab use based on its mechanism of action and the pathophysiology of such disorders. Key findings are presented by medical discipline, with respective indications discussed for which there is a scientific rationale for using rituximab based on the involvement of antibodies, antigens, T cells, and cytokines, all associated with B cell activity. By highlighting the underappreciated possibilities of rituximab repurposing through different lines of evidence demonstrating treatment benefit, we aim to inform healthcare professionals on its use and potential for future applications in over 200 B-cell mediated autoimmune diseases.

## Methodology

### Information sources for literature search

A literature search for on- and off-label uses of rituximab biosimilars was conducted between September 2021 and September 2022. This analysis started with a review of the Riximyo (rituximab) Product Monograph by Sandoz Canada Incorporated; its Regulatory Decision Summary and Summary Basis of Decision from Health Canada; the Highlights of Prescribing Information for RITUXAN (rituximab) by Biogen and Genentech; its Investigator’s Brochure, issued by Roche; its Biologics License Application; its FDA approval history; and the Medicare Coverage Database for off-label use of rituximab and rituximab biosimilars. Based on collected information, the methodology was broken down into four main sections: 1-establishing eligibility criteria, 2-search strategy and selection process in identifying (auto)immune diseases and rituximab use, 3-data collection process for defining key players in (auto)immunity, and 4-synthesis methods in categorizing disorders and depicting patent expiration (Table 1).

**Table 1:**
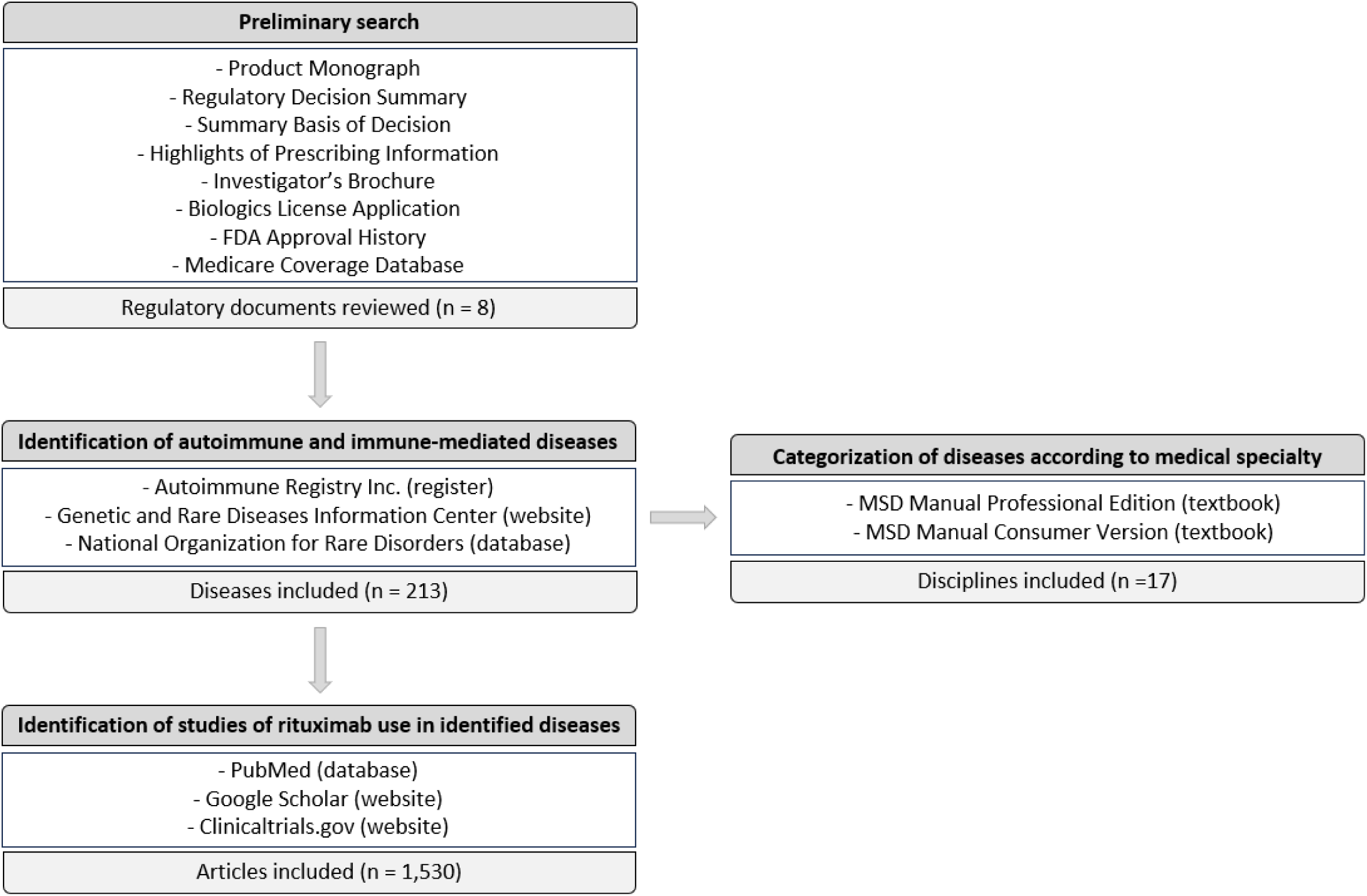
Flow chart of sytemic search strategy.

### Eligibility criteria

As approved (on-label) indications can vary from one regulatory body to the next, we defined off-label use according to Health Canada’s approval guidelines. Research articles published worldwide in any language or year were identified to recognize the extent of documented rituximab use. No limitations were imposed on the type or level of evidence gathered, which included anecdotal, observational, and experimental use. Any reported clinical benefit of rituximab in a given medical condition was included, regardless of study criteria, outcomes, or significance. If a particular autoimmune or immune-mediated disease was characterized by many complications and rituximab seemed to improve at least one of its manifestation, the study was included. We called this data set “perceived benefit” to encompass all lines of evidence, avoiding excluding any indication where rituximab biosimilars could be useful. Because there is much overlap and interchangeability in the literature between autoimmune and immune-mediated diseases, both were included in the analysis. Every condition identified whether entirely or partly autoimmune/immune-mediated was included, regardless of its cause or etiology. Autoinflammatory disorders were excluded. As our focus was the repurposing of rituximab from its approved use in hematologic cancers to off-label use in B cell-mediated autoimmune diseases, we excluded the discipline of oncology. Animal models and other experimental disease models were also excluded.

We included reports on adults worldwide, but only those without concomitant diseases. Studies involving children were not included unless a given disease arose in childhood and continued into adulthood. In all reports, rituximab was used as a single agent or as an add-on to an ongoing drug regimen that could not be terminated or replaced. The sole exceptions to this criterion were the use of premedication typically given prior to rituximab infusion to avoid or mitigate infusion-related reactions, and the use of concomitant drugs taken as part of an ongoing drug regimen where rituximab was added for further benefit or to taper or wean off current medication.

### Search strategy and selection process in identifying (auto)immune diseases and rituximab use

To gather a list of autoimmune and immune-mediated diseases, we consulted the Autoimmune Registry Inc., the Genetic and Rare Diseases Information Center Website (a program of the National Institutes of Health), and the National Organization for Rare Disorders’ Rare Disease Database. The PubMed database was searched using the keywords “autoimmune disease,” “immune-mediated disease,” “autoimmunity,” or “immunity,” and “Riximyo,” “Rituxan,” “rituximab,” “rituximab mechanism of action,” or “pathophysiology.” All diseases identified were also included as keywords in the PubMed search, followed by “pathophysiology” to confirm the involvement of (auto)immunity, and separately with “rituximab” to identify its use therein. If a PubMed search failed to provide information, Google Scholar was searched using the same keywords to locate scientific and medical peer-reviewed journals. If no results were posted anywhere, the Clinicaltrials.gov website was consulted to identify ongoing or recently completed trials involving rituximab to determine its use in a particular autoimmune or immune-mediated disease. We categorized the following two scenarios as “no perceived clinical benefit”: when studies or results on rituximab use/efficacy were pending, or when rituximab was used but follow-up or outcomes regarding its use were not reported. Overall, 1,530 articles met the established criteria, resulting in an extensive list of indications where rituximab has been or could be used (Supplementary Tables 1– 17).

### Data collection process for defining key players in (auto)immunity

Key immune cells or proteins were identified for every autoimmune and immune-mediated disease. We focused on B cell-mediated autoimmunity through (auto)antibodies, (self-)antigens, T lymphocytes, and inflammatory cytokines. If the articles initially gathered regarding disease pathophysiology did not specify the key players, we searched PubMed for additional information by combining the name of the disease with the following keywords: “B cells,” “B lymphocytes,” “B cell dysregulation,” “autoreactive B cells,” “autoreactive B lymphocytes,” “antibodies,” “autoantibodies,” “antigens,” “autoantigens,” “self-antigens,” “T cells,” “T lymphocytes,” “T cell involvement,” “autoreactive T cells,” “autoreactive T lymphocytes,” “cytokines,” “inflammatory cytokines,” “cytokine network,” and “cytokine imbalance” – the latter included either an increase in pro-inflammatory or a decrease in anti-inflammatory cytokines.

### Synthesis methods in categorizing disorders and depicting patent expiration

Where applicable, the Merck (MSD) Manuals (both the professional and consumer editions) were used to standardize and categorize the autoimmune and immune-mediated diseases identified. All potential off-label indications found were grouped by medical discipline. If a given disease or disorder did not appear in either of the Merck Manual searches, previously recorded data on the affected tissues or organ systems were used to classify it into the appropriate category.

To depict the impact of time and patent expiration on rituximab repurposing, the cumulative number of referenced articles in the current review was plotted relative to the year of publication. Using the 17 supplementary figures as a database encompassing 1,530 publications, articles from every medical specialty were included if they demonstrated a clinical benefit following rituximab treatment.

## Results

We identified 213 clinical indications that could benefit from off-label use of rituximab biosimilars across 17 medical specialties (rheumatology, neurology, dermatology, endocrinology, hematology, ophthalmology, gastroenterology, nephrology, immunology, cardiology, psychiatry, gynecology, pulmonology, hepatology, otolaryngology, urology, and infectiology). In the following sections, we describe in detail how rituximab has been used in these specialties, which are listed in descending order based on the number of B cell-mediated diseases identified. For each specialty, a table was generated to summarize the involvement of (auto)antibodies, (self-)antigens, T cells, and cytokines in the pathophysiology of all the selected clinical indications (Supplementary Tables 1–17). Moreover, the last column of each table indicates whether there are documented clinical benefits of using rituximab for each indication. The absence of clinical benefit was often due to the lack of rituximab repurposing for a given indication.

### Rheumatology

Rituximab repurposing began in 1999, after a case report describing clinical remission of RA in a patient with non-Hodgkin lymphoma treated with rituximab.^31^ Rituximab was approved as an RA treatment in 2006.^32^ Interestingly, we discovered that rituximab use has expanded to > 40 musculoskeletal and connective tissue disorders in rheumatology. These encompass joint and bursa, bone, and muscle and tendon disorders; autoimmune rheumatic disorders; vasculitides; and immunoglobulin G4-related disease (Supplementary Table 1). Mechanistically, B cells are crucial for tissue destruction across these indications, causing immune cells to infiltrate and accumulate in the synovium, juxta-articular regions, exocrine glands, and muscles, bones, and/or blood vessels, where they amplify immune and inflammatory responses. In addition to their local presence, these immune cells also end up in the systemic circulation, contributing to continuous immune activation and maintaining chronic disease. This translates into symptomatic manifestations and the perpetuation or worsening of the condition.

B cells play crucial roles in RA pathophysiology.^33–38^ They produce autoantibodies (rheumatoid factors and antibodies against citrullinated and carbamylated proteins) that bind self-antigens, contributing to immune complex formation and complement activation in and around the joints. These antibodies also activate T cells, which synthesize proinflammatory cytokines, promoting leukocyte infiltration of the joints, ectopic lymphoid center formation, angiogenesis, and synovial hyperplasia.^39^ Rituximab treatment dampens RA autoimmunity by depleting the source of these reactive autoantibodies.

Depleting B cells with rituximab diminished clinical symptoms by decreasing autoantibody concentrations, reducing proinflammatory cytokine levels, and lessening neutrophil extracellular trap release, which plays a role in autoantigen generation. This vicious cycle is at the core of autoimmune diseases, and rituximab’s ability to break it has extended its use to other indications in rheumatology.^40^ Although CD20^+^ B cell depletion varies among patients and may not necessarily be complete, the response to therapy has been positive across musculoskeletal and connective tissue disorders. Rituximab has yielded clinical benefits in 65% (26/40) of the identified autoimmune and immune-mediated diseases in rheumatology for which a rationale for its off-label use exists (Supplementary Table 1). Symptoms of inflammation are particularly alleviated;—patients feel less pain and body aches, experience less swelling, and ultimately suffer less structural joint and bone damage. This improvement in symptoms increases quality of life, with improved mobility and functional capacity (FIG. 2).

### Neurology

We identified 40 neurologic disorders using our research criteria, including demyelinating disorders; multiple sclerosis; autoimmune encephalitis; peripheral nervous system disorders (such as neuritis, neuropathies, as well as muscle cramps, spams, twitches, jerks, and involuntary movements); motor unit disorders and motor neuron diseases; neuromuscular junction disorders and disorders of neuromuscular transmission; autonomic nervous system disorders or autoimmune dysautonomia; sleep and wakefulness disorders; periodic limb movement disorders; movement and cerebellar disorders; central nervous system or brain and spinal cord disorders; pain; headaches and headache disorders such as migraines; delirium and dementia; tic disorders; and seizure disorders and autoimmune-related epilepsies (Supplementary Table 2). The notion that B cells could be involved in the pathophysiology of these neurologic disorders stemmed from the detection of antibodies in the cerebral spinal fluid (CSF) and/or at sites of injury of affected patients.^41^ Additional studies are needed to better understand the mechanistic involvement of B cells in the pathology of these indications. B cells secrete autoantibodies (from clonally expanded plasma cells in the CSF) and inflammatory cytokines, and act as autoimmune APCs. In addition to acting locally, they contribute to pathology from the periphery, causing and perpetuating disease symptoms. Although the symptoms associated with various neurological conditions are extremely heterogeneous, they all involve B cells entering, accumulating, and persisting in the central nervous system or its barriers.^41^ As such, halting disease progression requires eliminating these lymphocytes.

The most common neurological indication for rituximab repurposing is multiple sclerosis (MS), the most frequently occurring neuroimmunological disorder in young adults.^42–54^ Rituximab was the first anti-CD20 therapy used in MS, and is commonly prescribed off-label to treat different types of the condition.^55^ Depleting B cells with rituximab profoundly reduces clinical disease and signs of inflammation, as detected by magnetic resonance imaging scans. Patients experience less frequent relapses; benefit from a delayed disease progression; avoid the onset of certain expected symptoms altogether; and develop less gadolinium-enhancing lesions than untreated patients. This improves their quality of life, allowing them to independently perform their daily activities. Although these reported benefits are specific to MS, the same logic applies to the other identified neurological indications. To date, rituximab therapy has benefited 50% (20/40) of all autoimmune and immune-mediated diseases identified as neurological (Supplementary Table 2).

### Dermatology

We identified 23 dermatological disorders associated with B cell dysfunction (Supplementary Table 3). These include bullous diseases or intraepithelial autoimmune blistering dermatoses; psoriasis and scaling diseases; hypersensitivity and reactive skin disorders (dermatitis, dermatosis, hives or urticaria, and panniculitis); acne and related disorders, including autoinflammatory keratinization diseases; pigmentation disorders; and hair loss. The evolving understanding of these dermatological conditions has shifted the focus of recent studies to their underlying immune mechanisms.^56^ Many are mediated by B cells and their diverse functions, specifically autoantibody production (IgG or IgA), cytokine secretion, antigen presentation, and co-stimulatory effects.^56^ The most common symptoms include erythema, pruritus, hypo- or hyperpigmentation, xerosis or xeroderma, and fluid-filled pustules or blisters.^57^

In dermatology, the most well-known and successfully treated disorder with rituximab is PV, a chronic autoimmune disease characterized by the production of antibodies against desmogleins, a family of cellular adhesion proteins.^58–65^ Autoantibody-induced loss of adhesion between keratinocytes causes blisters and widespread erosions.^56^ Rituximab depletes these B cells, decreasing anti-desmoglein autoantibodies and desmoglein-specific T cells.^66^ Rituximab is effective in treating both untreated and previously treated moderate-to-severe PV. The treatment is well tolerated and significantly more effective at inducing prolonged remission than conventional therapy, without the side effects linked with steroid use. In case of relapse, repeated rituximab treatment is usually successful and is more effective when administered early in the disease course. Further studies may highlight the need for additional cycles of rituximab to maintain remission and establish the ideal dosage regimen.^58^ Coined a magic bullet for PV, rituximab is now approved for its treatment by the FDA but is still prescribed off-label in Canada.

Beyond PV, clinical and/or laboratory improvements have been observed in 65% (15/23) of identified autoimmune and immune-mediated dermatological disorders for which there is a rationale for off-label rituximab use (Supplementary Table 3). Treated patients experience a prolonged clinical remission, typically correlating with sharp decreases in lymphocytes and antibody titers. Rituximab provides relief against skin itchiness, redness, swelling, dryness, and/or eruptions, as measured by visual analog scales and disease severity indexes.

### Endocrinology

Our criteria identified 20 endocrine and metabolic disorders characterized by the aberrant involvement of B cells, resulting in the secretion of pathogenic IgG antibodies that attack components of the hypothalamic-pituitary-adrenal axis and related glands (Supplementary Table 4). These included polyglandular deficiency syndromes, adrenal disorders, and adrenalitis, including different types of adrenal insufficiencies; thyroid disorders and thyroiditis; parathyroid disorders and parathyroiditis; pituitary disorders and multiple variants of hypophysitis; diabetes mellitus and other insulin syndromes and autoimmune forms of hypoglycemia; dyslipidemia; and protein aggregation and deposition.

Graves’ disease (GD) is a common endocrine disorder with multiple associated complications. It is a thyroid autoimmune disorder in which stimulatory anti-thyrotropin receptor autoantibodies (TRAb) produced by reactive B cells induce hyperthyroidism.^67–76^ Patients suffering from GD are also affected by Graves’ orbitopathy (GO), its main extra-thyroidal manifestation. GO is characterized by the inflammation and remodeling of orbital tissues in response to antibody attacks and dysregulated cytokine expression, including chemoattractants.^77^ GO responds effectively to rituximab when used early in the disease course.^67^ Rituximab relieves the disease symptoms relatively quickly, exerting its clinical effects only a few weeks after the first infusion.^67^ Treated patients have decreased clinical activity scores (an index of disease severity) and relief from proptosis and diplopia, and increased quality of life. These effects are likely due to significantly reduced serum TRAb levels and decreased peripheral B cells.^67^

To date, 45% (9/20) of all autoimmune and immune-mediated diseases identified in endocrinology that have a rationale for off-label rituximab therapy have benefited from its use (Supplementary Table 4).

### Hematology

Based on our search criteria, we identified 15 B cell-mediated clinical indications in hematology (Supplementary Table 5). Several types of cytopenias fall under this category, affecting red blood cells (erythroid precursors and erythrocytes), white blood cells (neutrophils and lymphocytes), and/or platelets (thrombocytes), as well as hemolytic anemias, anemias caused by deficient or decreased erythropoiesis, leukopenias (such as neutropenia and lymphocytopenia), thrombocytopenias and platelet dysfunction, coagulation and thrombotic disorders, lymph node affections, and plasma cell disorders.

The aberrant involvement of B cells in hematological diseases seems more intuitive than in other medical disciplines, as B cells are a main component of blood. B cells can produce autoantibodies against various blood constituents, from precursors to mature blood cells, causing a heterogeneous group of conditions called autoimmune cytopenias, which can be treated with rituximab.^78–93^ In addition, immune thrombocytopenic purpura (ITP) is mediated by platelet or thrombocyte autoantibodies.^94–101^ These autoantibodies often target the two most prevalent receptors on the platelet surface (the GPIb/IX complex (von Willebrand factor receptor) and the GPIIb/IIIa receptor (collagen/fibrinogen receptor), reducing the ability of the platelets to aggregate.^93^ As a result, patients present with variable degrees of mucocutaneous bleeding. Rituximab has been widely used off-label to treat ITP as it raises platelet count by inhibiting their destruction. In doing so, it decreases symptoms such as petechiae, purpura, hematomas, gum and nose bleeding, blood in urine or stools, as well as extreme tiredness.^102^ Furthermore, the administration of rituximab early in the disease course has also showed to improve relapse-free survival.^103^ We found that 87% (13/15) of autoimmune and immune-mediated hematological conditions with a rationale for off-label rituximab use benefited from the therapy (Supplementary Table 5).

### Ophthalmology

Our analysis identified 13 eye disorders linked to B cells (Supplementary Table 6). These comprised several forms of autoimmune uveitis and related disorders; corneal disorders; conjunctival and scleral disorders; retinal and optic nerve disorders; glaucoma; idiopathic orbital inflammation; and lacrimal and orbital diseases. Although how B cells are involved in these disorders is not entirely understood, they are instrumental for autoimmune tissue destruction in and around the eye. In addition to secreting autoantibodies that directly induce cell lysis and are involved in antigen-antibody immune complex deposition into the ocular tissue, B cells promote intraocular inflammation by presenting antigens to T cells, producing many inflammatory cytokines, and supporting T cell survival.^104,105^ Patients with active eye inflammation have significantly more infiltrating B cells in their ocular tissues and higher levels of B cell-associated cytokines in their aqueous humors than healthy controls.

Uveitis, a diverse group of intraocular inflammatory diseases, is the autoimmune eye disorder most commonly treated with rituximab.^105^ It produces a positive therapeutic response by depleting the CD20^+^ B cells that attack various segments of the eye, including disease improvement and remission, which translates into improved visual acuity.^104,106–120^

Off-label rituximab has produced clinical benefits in 62% (8/13) of all B cell-mediated autoimmune and immune-mediated eye diseases (Supplementary Table 6), improving the vision of patients suffering from all ophthalmological disorders tested. Based on the success of rituximab against GO, it may be interesting to induce B cell depletion with rituximab shortly after diagnosis instead of reserving it as a second-line therapy.^74^

### Gastroenterology

Nine B cell-mediated gastrointestinal (GI) disorders were identified, including swallowing disorders, multiple affections of GI tract components and accessory digestive organs (*i.e.*, esophagitis, pancreatitis, gastritis, enteropathies, and polyposis), and many forms of colitis or inflammatory bowel disease (IBD), which affect the intestines and rectum (Supplementary Table 7).

In contrast to other categories covered thus far, rituximab use in gastroenterology is limited. Rituximab administered for non-GI indications can induce GI disorders. Recent reports have linked rituximab with the development of *de novo* colitis, a type of IBD. Other reported rituximab-associated GI toxicities include diarrhea and bowel perforation.^121^ These side effects could result from the infusion itself, suggesting that using rituximab therapy for GI disorders should be approached with caution. Nonetheless, B cells are heavily involved in the pathophysiology of these diseases. B cells produce IgG in the gut, which contributes to intestinal inflammation in IBD.^122,123^ One of the few GI disorders for which rituximab showed a favorable response, including safety and efficacy, is autoimmune pancreatitis.^124–132^ Patients afflicted with this disease experience remission, and consequently, relief from severe abdominal pain after rituximab treatment. Overall, rituximab provided clinical benefit in 22% (2/9) of the identified autoimmune and immune-mediated GI diseases whose pathophysiologies indicate a rationale for its repurposing (Supplementary Table 7).

### Nephrology

We identified seven B cell-mediated kidney disorders, including multiple glomerular disorders (nephritides, nephropathies, glomerulopathies, and podocytopathies) and tubulointerstitial diseases (Supplementary Table 8). In contrast to gastroenterology, where rituximab use is limited, its repurposing in nephrology is very promising as 86% (6/7) of the identified indications showed a favorable response to off-label therapy (Supplementary Table 8). Accordingly, rituximab use in nephrology is increasing.^133,134^ B cell clonal expansion has been observed in the blood and/or renal tissue of patients with glomerulonephritis, which rituximab targets.^135–140^ Glomerulonephritis (*i.e*., inflammation of the glomeruli) is characterized by pathogenic fibril deposition within damaged kidney components as a consequence of autoimmune processes governed by B cells.^141^ Rituximab-mediated B cell depletion alleviates many disease symptoms, including proteinuria. Importantly, kidney function is preserved and even improved after treatment is completed.^137,142–146^

### Immunology

Seven B cell-mediated immune and allergic disorders were identified (Supplementary Table 9). These include immunodeficiency disorders or primary immune deficiency diseases, inborn errors of immunity, transplantation complications, allergic and atopic disorders, and other immune hypersensitivity disorders. As in hematology, B cell involvement in immunologic disorders seems quite intuitive, as they are essential for the adaptive immune response, humoral immunity, and antibody production. The activation of autoreactive B cells drives the pathogenic processes of autoimmune diseases.^147^ Therefore, B cell targeting has become an effective treatment modality for immune and allergic disorders.^148^ In particular, rituximab is a key medication used in antibody-mediated rejection (AMR), a common and potentially devastating sequela of allograft rejection associated with graft loss after transplantation. AMR is a leading cause of morbidity and mortality following transplant surgery.^149^ In AMR, B cells from the donor produce antibodies against antigens expressed on recipient organ cells.^150–160^ Rituximab administration before transplantation, with or without follow-up maintenance doses, improved AMR and graft survival rates in patients undergoing kidney transplantation.^161^ Similarly, rituximab may also improve clinical responses and quality of life in patients who develop steroid-refractory chronic graft-versus-host disease after allogeneic stem cell transplantation.^162–166^ Of the identified autoimmune and immune-mediated diseases for which rituximab was used off-label, 86% (6/7) of them have benefited from rituximab therapy (Supplementary Table 9).

### Cardiology

We identified seven cardiovascular disorders characterized by the inflammation of various cardiac tissues (myocarditis, pericarditis, endocarditis, arteriosclerosis, peripheral arterial disorders, and diseases of the aorta plus its branches; Supplementary Table 10). As rituximab infusion is associated with adverse cardiac side effects (including hypotension, hypoxia, myocardial infarction, arrhythmias, cardiogenic shock, and dilated cardiomyopathy), its use in cardiology may seem counterintuitive. However, considering the marked involvement of B cells in cardiac disease pathophysiology, rituximab therapy can be beneficial, especially for patients with heart failure.^167^ Heart-reactive autoantibodies in the sera of affected patients interfere with cardiomyocyte function by targeting diverse antigens, including α- and β-myosin heavy-chain isoforms, sarcolemma proteins, mitochondrial enzymes, and beta-adrenergic and muscarinic receptors.^168–170^ B cells also activate fibroblasts and a variety of immune cells (T cells, neutrophils, macrophages). Altogether, these activated cells promote the development of cardiac hypertrophy, inflammation, maladaptive tissue remodeling, and fibrosis, which ultimately lead to heart failure. Because of their active involvement in the disease, rituximab has been used to target B cells, leading to improved left ventricular ejection fractions^167,171^ and better overall quality of life for patients with autoimmune and immune-mediated cardiovascular diseases. We found that 57% (4/7) of these indications have benefited from rituximab repurposing (Supplementary Table 10).^168,170,172–183^

### Pulmonology

We identified five pulmonary disorders mediated by B cell, including idiopathic pulmonary fibrosis (IPF), non-IPF interstitial lung diseases (ILDs), asthma and related disorders, diffuse alveolar or pulmonary hemorrhage, and sarcoidosis, 60% (3/5) of which have benefited from rituximab repurposing (Supplementary Table 11). ILDs are a group of heterogeneous disorders characterized by lung inflammation and scarring, with alveolar septal thickening, fibroblast proliferation, collagen deposition, and ultimately pulmonary fibrosis. ILDs with no clear etiology are typically autoimmune or immune-mediated and are termed “idiopathic interstitial pneumonias.” Affected patients suffer from cough, dyspnea, and shortness of breath due to the abnormal accumulation of inflammatory blood cells. White blood cells (B cells and macrophages) and fluid accumulate in the lung tissue, which cause and perpetuate inflammation.^184–191^ If left untreated, scar tissue replaces healthy lung tissue and the alveoli are progressively destroyed, ultimately leading to pulmonary fibrosis. Various degrees of B cell infiltration occur in different forms of ILD.^192^ By eliminating B cells, rituximab improves both IPF and non-IPF ILD, improving patients’ pulmonary function and quality of life.^188,193–198^

### Hepatology

We identified four B cell-mediated hepatic and biliary disorders, including autoimmune hepatitis (AIH) different stages of liver scarring such as fibrosis and cirrhosis, and gallbladder and bile duct disorders including cholangitis (Supplementary Table 12). B cells are integral to autoimmune liver pathogenesis,^199^ promoting autoimmunity by presenting antigens to T cells, producing autoantibodies, and generating inflammatory cytokines.^200–205^ AIH is clinically characterized by the presence of antibodies against liver autoantigens, hypergammaglobulinemia, and a lymphoplasmocytic infiltrate with interface activity (by liver histology).^199^ AIH can result in hepatic fibrosis, progress to cirrhosis, and eventually require liver transplantation. Rituximab is effective against AIH, causing sustained improvements in serum liver enzymes and an absence of clinical disease in the first few years after therapy.^201,206,207^ Rituximab treatment also helps reduce the burden of corticosteroid use and associated side effects.^207^ As such, patients experience a better quality of life, and can delay or completely avoid liver transplantation. However, the use of rituximab in hepatology should be approached with caution, as several studies have reported liver toxicity or liver failure secondary to reactivation of hepatitis B following rituximab infusion (also known as “rituximab-induced autoimmune hepatitis”). Although rituximab may not replace the current standard of care, it could prove useful in specific cases by controlling disease flare-ups.^208^ Among all the identified autoimmune and immune-mediated hepatic diseases that could be treated with off-label rituximab, 50% (2/4) responded favorably (Supplementary Table 12).

### Other disciplines

Finally, we collated data on five other medical disciplines with single or no indications shown to benefit from rituximab therapy: psychiatry, gynecology, otolaryngology, urology, and infectiology (Supplementary Tables 13–17). Nevertheless, B cells are crucial for the self-tissue damage at the heart of their pathophysiologies, through the same mechanisms described for other specialties (FIG. 2). However, despite a rationale for repurposing rituximab to improve their clinical symptoms, rituximab use has not been reported for many of these B cell-driven indications, with the exception of pemphigoid gestationis^209–215^ (gynecology) and autoimmune inner ear disease^216–219^ (otolaryngology). Therefore, these lists of autoimmune diseases contain promising candidates for future rituximab repurposing.

### Repurposing trends over time

Lastly, we examined rituximab repurposing over time, relying on our curated database of 1,530 references (Supplementary Tables 1–17). As a metric of its repurposing in patients, we calculated annual cumulative numbers of referenced articles per medical specialty. Of the 17 medical specialties studied, 13 showed favorable clinical responses to rituximab (FIG. 3). Interestingly, rituximab repurposing accelerated approximately 10 years after its regulatory approval for hematological malignancies. Rheumatology and neurology had the highest repurposing rates, followed by dermatology. Moreover, rituximab repurposing tended to increase around 2018, when its US patent expired, suggesting that access to less expensive biosimilars may stimulate its repurposing for a vast number of indications, including rare or orphan diseases.

**Fig. 3:**
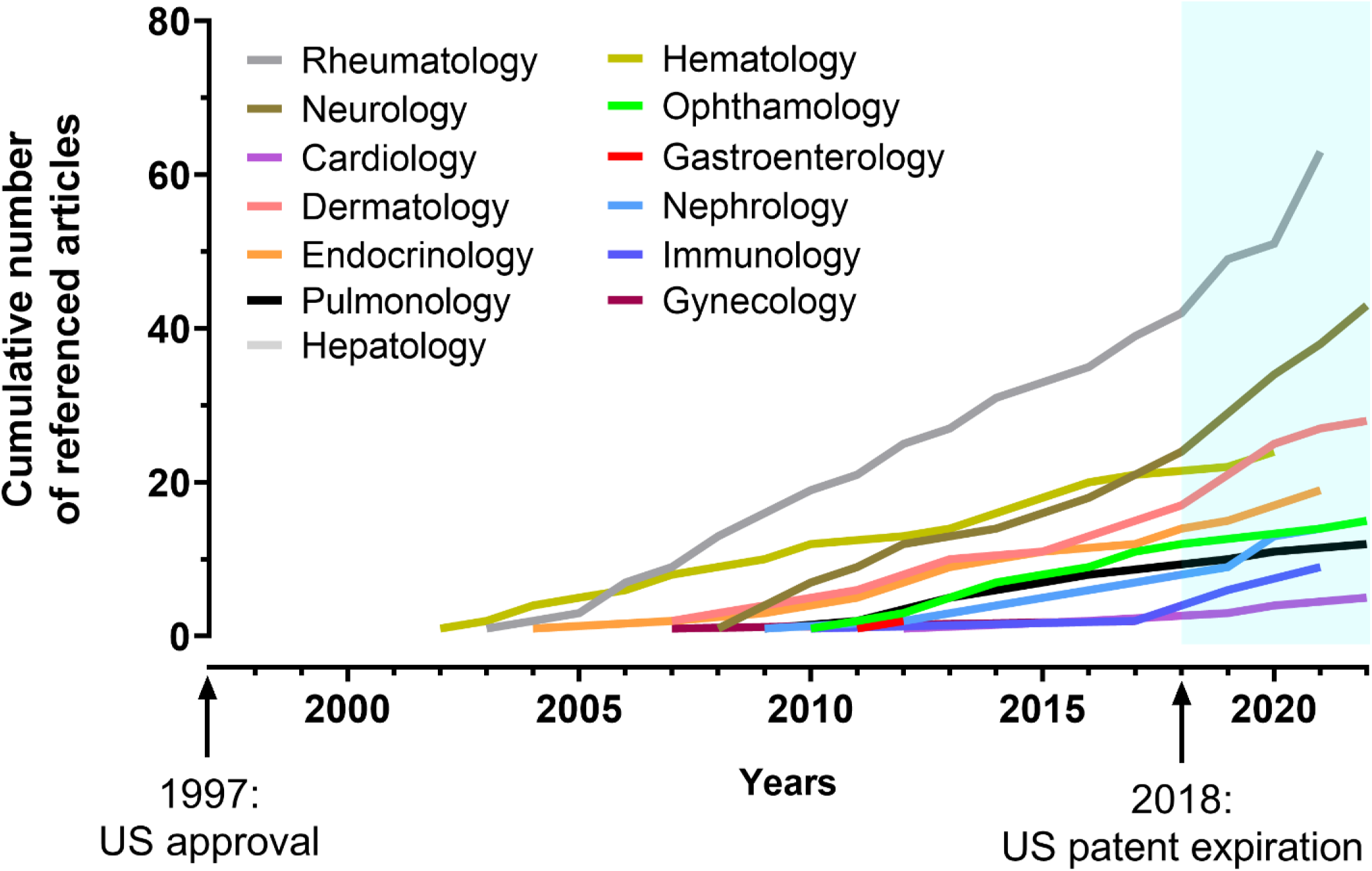
Trends in rituximab use over time, from approval to patent expiration and beyond. To visualize the impacts of time and patent expiration on rituximab repurposing, the 1,530 articles referenced in this review were plotted based on their year of publication. The years of rituximab’s approval and expiration are indicated.

## Discussion

Our comprehensive analysis of 213 autoimmune and immune-mediated diseases showed that rituximab has been extensively prescribed off-label therein, producing documented clinical benefit in 116 indications across 17 medical specialties. As B cells play central roles in a myriad of autoimmune diseases, we also highlighted the repurposing potential of rituximab in naïve indications or in those where further testing is required to obtain results of therapeutic outcome. Figure 4 shows the impressive number of indications demonstrating a clinical benefit alongside a number of potential indications for rituximab use in B cell-mediated diseases. Altogether, our work underlines the promise of rituximab repurposing, due to its impacts on CD20^+^ B cell-mediated autoimmune diseases addressing in some cases urgent unmet clinical needs, particularly in cases that are refractory or intolerant to current standards of care.

**Fig 4:**
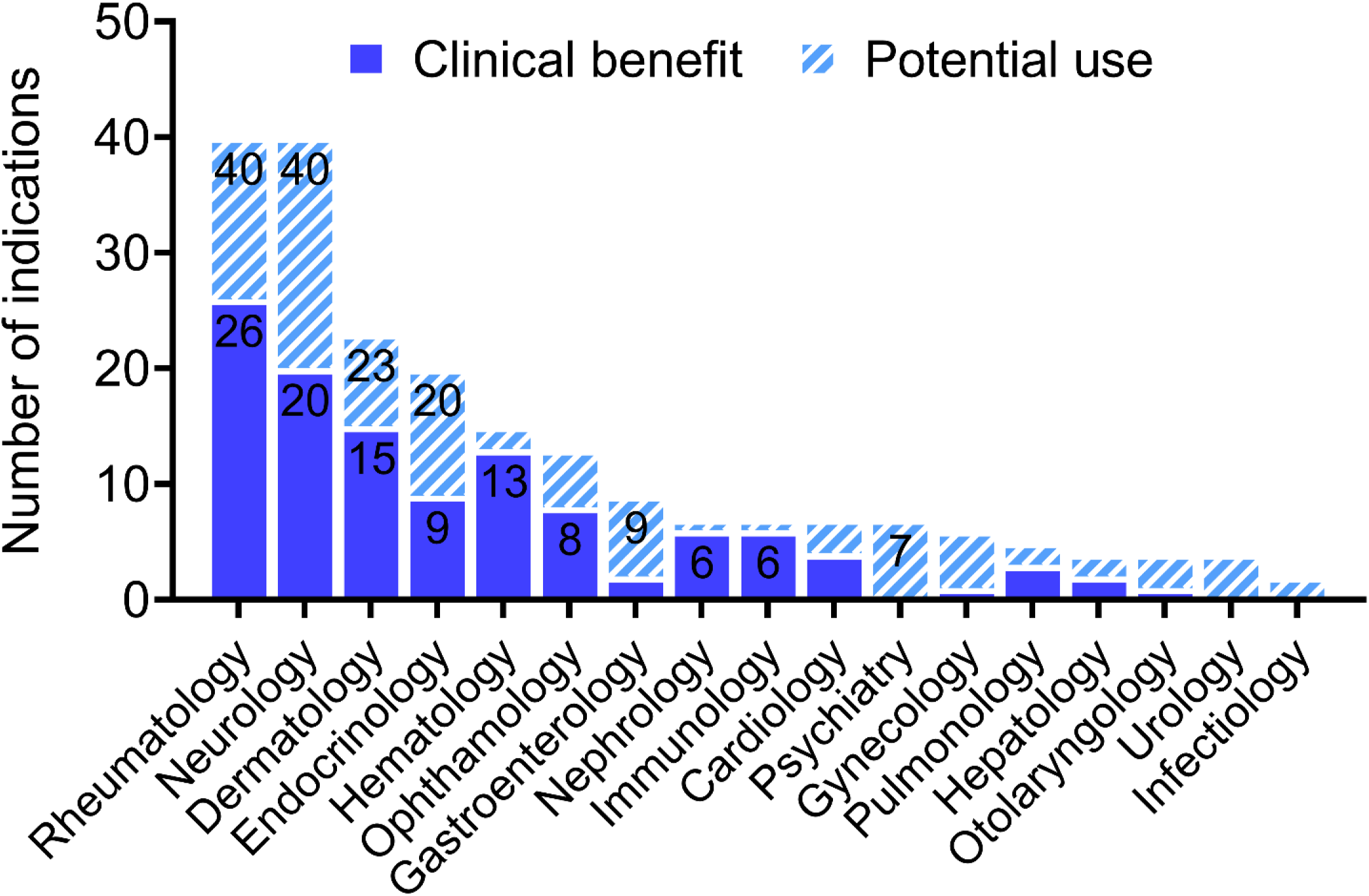
Rituximab repurposing across 17 medical specialties. The number of indications per medical specialty that may benefit from rituximab repurposing, based on its mechanism of action and the pathophysiology of the autoimmune disease. This is plotted alongside the number of indications for which rituximab has been used with demonstrated clinical benefit.

Although the underlying pathophysiologies of the autoimmune diseases for which rituximab has been repurposed are not fully elucidated or necessarily similar, B cells are their common link. The pathological cascade follows a comparable sequence, *i.e.*, loss of self-tolerance causes B cells to produce autoantibodies, present antigens, and activate T lymphocytes, which in turn secrete inflammatory cytokines that contribute to disease.^11^ As such, eliminating CD20^+^ B cells restores immune tolerance and rewires the immune system such that self-proteins are no longer perceived as foreign.^220^ As this review demonstrates, rituximab could be repurposed for numerous autoimmune and immune-mediated diseases across various medical specialties.^32^

Given the high failure rate in developing new medicines, repurposing drugs to treat diseases with similar molecular mechanisms is extremely attractive, de-risking the drug development path by lowering global costs and shortening the development time. The cost to develop a new molecular compound can reach 2–3 billion USD, while drug repurposing is significantly less expensive, with an estimated average of 300 million USD. The case for biologic repurposing is even more interesting, as their costs and development times are generally even longer than those for small molecules.^13^ In the history of medicine, drug repurposing has mainly occurred by serendipity, with a small molecule typically repurposed for a single indication or, at best, a couple of indications. The greatest advantage of rituximab repurposing is its applicability to more than 100 different indications. In addition to its approved uses (hematological malignancies, RA, granulomatosis with polyangiitis, and microscopic polyangiitis), a significant portion of sales of rituximab biosimilars is off-label. Rituximab’s patent expiration at the end of the 1990s may have prompted pharmaceutical companies to refrain from investing in novel clinical trials, particularly for rare diseases. However, the success of its repurposing has also stimulated the industry of biosimilars.

As with any biologic product, research and development costs are steep, making rituximab difficult to afford without proper health insurance. Fortunately, since its patent expiration, rituximab biosimilars such as Riximyo have made treatment more affordable and accessible.

Once a drug has demonstrated either off-target effects or a newly identified on-target effect (as in the case of rituximab), it may be prescribed as an off-label treatment or tested in clinical trials to expand its approved range of indications. Physicians are allowed to use their knowledge, the scientific literature, and even anecdotal case reports to prescribe drugs that may relieve patient symptoms.^221^ Importantly, in the context of rare diseases, which are often neglected by pharmaceutical industries due to the low chance of a return on the investment, drug repurposing is an appealing solution. Off-label use avoids the need to obtain regulatory approval through clinical trials, which is not practical for small patient populations. Rituximab is often repurposed for B cell-mediated diseases because, in many cases, the approved treatments are ineffective. However, although many studies and case reports support rituximab repurposing for a wide range of clinical indications, the concern, which may limit its repurposing, may be associated with the treatment-induced adverse effects for a drug without official approval.

B cells are at the core of immunity and depleting them comes with a cost; therefore, it is important to evaluate the risks and benefits of rituximab therapy on a case-by-case basis. The immunosuppression caused by rituximab is a double-edged sword: it beneficially wipes out autoreactive B cells but depletes important blood cells, thereby inducing side effects that can require medical attention. Immunosuppression makes patients prone to infections, and they must be closely monitored during and after treatment. The depletion of circulating B cells after rituximab administration is rapid (within 1 week) and profound, with peripheral blood B cell counts remaining low for at least 6–12 months following therapy. The repopulating pool is dominated by immature B cells, while memory B cell recovery is slow and occurs after 2 years.^222,223^

Overall, this review gathered the documented clinical benefits of rituximab repurposing and well-established data regarding disease pathophysiology, providing a scientific rationale for using rituximab to treat autoimmune and immune-mediated disorders across different medical disciplines. Although many of the cited studies highlight the therapeutic benefits of rituximab in indications other than those originally approved, these small case reports and small-scale clinical studies did not reach the stage of randomized clinical trials. Therefore, even though these studies support the clinical benefits of rituximab repurposing, further studies are warranted to obtain a better understanding of its efficacy and safety across different indications—in larger patient cohorts, when possible. The economic burden of this biological therapy likely also contributes to the limited number of clinical studies. Ultimately, the decision to repurpose a drug depends on shared knowledge and available clinical observations, the personal experience of the treating physician, and the patient’s willingness to try a medication off-label.

## Conclusion

In summary, rituximab treatment appeared to be beneficial in 54% (116/213) of autoimmune and immune-mediated diseases identified across all 17 medical specialties. This finding highlights the need for future studies on the remaining rituximab-naïve indications that could potentially benefit from this therapy. In addition, for the 116 diseases that have already benefited from rituximab repurposing, larger studies are needed to confirm its efficacy or establish its long-term safety, in addition to establishing the optimal drug regimens for each specific indication. If the trend of rituximab repurposing continues to grow (FIG. 3), the combination of more scientific knowledge on B cell involvement in immune disease and a cost reduction of biosimilars will stimulate off-label use in the future.

Several well-known drugs have been successfully repurposed for 1–2 additional indications, including sildenafil, thalidomide, and celecoxib.^32^ Rituximab’s number of indications (116) and global sales (in the billions) greatly exceed those of other repurposed drugs.^32^ Therefore, rituximab is the most exemplary case of the value of drug repurposing since the concept emerged in the early 2000s.^224^ This review serves as a reference database that provides a rationale for using rituximab to treat patients afflicted with B cell-mediated diseases.

## Data Availability

All data produced in the present work are contained in the manuscript

## Acknowledgements

N.J.-M.R. holds a Junior 2 Research Scholar award from the Fonds de Recherche du Québec en Santé.

## Author contribution statement

A.M. collected the data and performed the analysis. A.M., G.R., and N.J.-M.R. wrote the manuscript.

## Acknowledgements

N.J.-M.R. holds a research scholar Junior 2 award from the Fonds de Recherche du Québec en Santé. We thank High-Fidelity Science Communications for manuscript editing.

