## Supplementary Tables 1-17 for "Repurposing of rituximab biosimilars to treat B cell mediated autoimmune diseases"

The 17 supplementary tables summarize how the immune system impacts medical indications in various specialties. A checkmark (✓) indicates a documented link between an indication and antibodies, antigens, T cells, and/or cytokines and a reported benefit from rituximab. A minus (–) indicates that no documented link was found during our literature search. References are listed for each indication.

**Supplementary Table 1: Musculoskeletal, and connective tissue disorders mediated by B cells**

Involvement of B cells in the pathophysiology of autoimmune or immune-mediated rheumatological, musculoskeletal, and connective tissue disorders through Ab-dependent and Ab-independent mechanisms, as supported by the presence of (auto)antibodies, (self)antigens, T cells, and/or cytokines. Reported benefit of rituximab is also noted for each indication. *Abbreviations*: SAPHO, synovitis, acne, pustulosis, hyperostosis, osteitis; IMNM, immune-mediated necrotizing myopathy; PM, polymyositis; DM, dermatomyositis; IBM, inclusion body myositis; ASS, antisynthetase syndrome.

| # | Indication | Antibodies | Antigens | T cells | Cytokines | Documented rituximab benefit | References |
|---|---|---|---|---|---|---|---|
| 1 | Rheumatoid arthritis | ✓ | ✓ | ✓ | ✓ | ✓ | 1-10 |
| 2 | Still disease (adult and juvenile) | ✓ | ✓ | ✓ | ✓ | ✓ | 11-20 |
| 3 | Ankylosing spondylitis | ✓ | ✓ | ✓ | ✓ | ✓ | 21-23 |
| 4 | Psoriatic arthritis | ✓ | ✓ | ✓ | ✓ | ✓ | 24-31 |
| 5 | Reactive arthritis | ✓ | ✓ | ✓ | ✓ | – | 32-39 |
| 6 | Osteoarthritis | ✓ | ✓ | ✓ | ✓ | – | 40-48 |
| 7 | Gout | ✓ | ✓ | ✓ | ✓ | – | 49-54 |
| 8 | Palindromic rheumatism | ✓ | ✓ | ✓ | ✓ | ✓ | 55-61 |
| 9 | Osteoporosis | ✓ | ✓ | ✓ | ✓ | – | 62-64 |
| 10 | SAPHO syndrome | ✓ | ✓ | ✓ | ✓ | – | 65-72 |
| 11 | Systemic lupus erythematosus | ✓ | ✓ | ✓ | ✓ | ✓ | 73-77 |
| 12 | Sjögren syndrome | ✓ | ✓ | ✓ | ✓ | ✓ | 78-86 |
| 13 | Systemic sclerosis | ✓ | ✓ | ✓ | ✓ | ✓ | 87-95 |
| 14 | Morphea | ✓ | ✓ | ✓ | ✓ | – | 87, 96-100 |
| 15 | Eosinophilic fasciitis | ✓ | ✓ | ✓ | ✓ | ✓ | 101-105 |
| 16 | Relapsing polychondritis | ✓ | ✓ | ✓ | ✓ | ✓ | 106-109 |
| 17 | Mixed connective tissue disease | ✓ | ✓ | ✓ | ✓ | ✓ | 110-115 |
| 18 | Undifferentiated connective tissue disease | ✓ | ✓ | ✓ | ✓ | – | 116-121 |
| 19 | Autoimmune myositis (IMNM, PM, DM, IBM, ASS) | ✓ | ✓ | ✓ | ✓ | ✓ | 122-140 |
| 20 | Acute rheumatic fever | ✓ | ✓ | ✓ | ✓ | – | 141-146 |
| 21 | Fibromyalgia | ✓ | ✓ | ✓ | ✓ | – | 147-149 |
| 22 | Parry-Romberg syndrome | ✓ | ✓ | ✓ | – | ✓ | 150-155 |
| 23 | Granulomatosis with polyangiitis | ✓ | ✓ | ✓ | ✓ | ✓ | 156-168 |
| 24 | Microscopic polyangiitis | ✓ | ✓ | ✓ | ✓ | ✓ | 168-176 |
| 25 | Eosinophilic granulomatosis with polyangiitis | ✓ | ✓ | ✓ | ✓ | ✓ | 177-184 |

| | | | | | | | |
|---|---|---|---|---|---|---|---|
| 26 | Immunoglobulin A–associated vasculitis | √ | √ | √ | √ | √ | 185-193 |
| 27 | Cryoglobulinemia | √ | √ | √ | √ | √ | 194-205 |
| 28 | Cutaneous vasculitis | √ | √ | √ | √ | √ | 206-210 |
| 29 | Polyarteritis nodosa | √ | √ | √ | √ | √ | 211-217 |
| 30 | Kawasaki disease | √ | √ | √ | √ | √ | 218-227 |
| 31 | Behçet disease | √ | √ | √ | √ | √ | 228-236 |
| 32 | Takayasu arteritis | √ | √ | √ | √ | √ | 237-246 |
| 33 | Giant cell arteritis | √ | √ | √ | √ | – | 242,247-250 |
| 34 | Polymyalgia rheumatica | √ | √ | √ | √ | – | 247,251-253 |
| 35 | Susac syndrome | √ | √ | √ | √ | – | 254-261 |
| 36 | Sneddon syndrome | √ | √ | √ | √ | – | 262-274 |
| 37 | Malignant atrophic papulosis | √ | √ | √ | √ | – | 275-281 |
| 38 | Immunoglobulin G4-related disease | √ | √ | √ | √ | √ | 282-293 |
| 39 | Felty syndrome | √ | √ | √ | √ | √ | 294-300 |
| 40 | Schnitzler syndrome | √ | √ | √ | √ | √ | 301-309 |

**Supplementary Table 2: Neurological disorders mediated by B cells**

Involvement of B cells in the pathophysiology of autoimmune or immune-mediated diseases in neurology through Ab-dependent and Ab-independent mechanisms, as supported by the presence of (auto)antibodies, (self)antigens, T cells, and/or cytokines. Reported benefit of rituximab is also noted for each indication. *Abbreviations*: MOG, myelin oligodendrocyte glycoprotein.

| # | Indication | Antibodies | Antigens | T cells | Cytokines | Documented rituximab benefit | References |
|---|---|---|---|---|---|---|---|
| 41 | Multiple sclerosis & clinically isolated syndrome | ✓ | ✓ | ✓ | ✓ | ✓ | 310-332 |
| 42 | Baló concentric sclerosis & myelinoclastic diffuse sclerosis | ✓ | – | ✓ | – | – | 333-336 |
| 43 | Neuromyelitis optica | ✓ | ✓ | ✓ | ✓ | ✓ | 337-342 |
| 44 | Autoimmune encephalitis | ✓ | ✓ | ✓ | ✓ | ✓ | 343-358 |
| 45 | Acute disseminated encephalomyelitis | ✓ | ✓ | ✓ | ✓ | ✓ | 359-367 |
| 46 | Acute hemorrhagic leukoencephalitis | ✓ | ✓ | ✓ | ✓ | – | 368,369 |
| 47 | Rasmussen encephalitis | ✓ | ✓ | ✓ | ✓ | ✓ | 353, 370-378 |
| 48 | Bickerstaff brainstem encephalitis | ✓ | ✓ | – | – | ✓ | 379-383 |
| 49 | Chronic inflammatory demyelinating polyneuropathy | ✓ | ✓ | ✓ | ✓ | ✓ | 384-391 |
| 50 | Myasthenia gravis | ✓ | ✓ | ✓ | ✓ | ✓ | 392-399 |
| 51 | Parsonage-Turner syndrome | ✓ | ✓ | ✓ | ✓ | – | 400-404 |
| 52 | Guillain-Barré syndrome & Miller Fisher syndrome | ✓ | ✓ | ✓ | ✓ | – | 405-411 |
| 53 | Lambert-Eaton myasthenic syndrome | ✓ | ✓ | ✓ | – | ✓ | 412-419 |
| 54 | Stiff-person syndrome | ✓ | ✓ | ✓ | ✓ | ✓ | 420-432 |
| 55 | Peripheral nerve hyperexcitability syndromes | ✓ | ✓ | – | – | ✓ | 433-446 |
| 56 | Small fiber neuropathy | ✓ | ✓ | – | ✓ | – | 447-451 |
| 57 | Multifocal motor neuropathy | ✓ | ✓ | ✓ | ✓ | ✓ | 452-460 |
| 58 | Amyotrophic lateral sclerosis | ✓ | ✓ | ✓ | ✓ | – | 461-469 |
| 59 | Satoyoshi syndrome | ✓ | ✓ | – | – | – | 470, 471 |
| 60 | Autoimmune autonomic ganglionopathy | ✓ | ✓ | – | – | ✓ | 472-477 |
| 61 | Autoimmune gastrointestinal dysmotility | ✓ | ✓ | – | – | – | 478-480 |
| 62 | Narcolepsy type 1 | ✓ | ✓ | ✓ | ✓ | ✓ | 481-488 |
| 63 | Kleine-Levin syndrome | ✓ | ✓ | ✓ | ✓ | – | 489-492 |
| 64 | Restless legs syndrome | ✓ | ✓ | ✓ | ✓ | – | 493-497 |
| 65 | Chronic fatigue syndrome | ✓ | ✓ | ✓ | ✓ | ✓ | 498-504 |
| 66 | Huntington disease | ✓ | ✓ | ✓ | ✓ | – | 505-509 |
| 67 | Parkinson disease | ✓ | ✓ | ✓ | ✓ | – | 510-514 |
| 68 | Autoimmune cerebellar ataxia | ✓ | ✓ | ✓ | ✓ | ✓ | 515-532 |

| | | | | | | | |
|---|---|---|---|---|---|---|---|
| 69 | Opsoclonus-myoclonus syndrome | √ | √ | √ | √ | √ | 533-541 |
| 70 | Sydenham chorea | √ | √ | √ | √ | – | 542-547 |
| 71 | Transverse myelitis | √ | √ | √ | √ | – | 548-553 |
| 72 | MOG antibody-associated disease | √ | √ | √ | √ | √ | 554-558 |
| 73 | Autoimmune glial fibrillary acidic protein astrocytopathy | √ | √ | √ | √ | √ | 559-568 |
| 74 | Complex regional pain syndrome | √ | √ | √ | √ | – | 569-578 |
| 75 | Moyamoya disease | √ | √ | √ | √ | – | 579-586 |
| 76 | Tolosa-Hunt syndrome | √ | √ | √ | – | – | 587,588 |
| 77 | Autoimmune dementia | √ | √ | √ | √ | – | 589-597 |
| 78 | Creutzfeldt-Jakob disease | √ | √ | √ | √ | – | 598-604 |
| 79 | Tourette syndrome | √ | √ | √ | √ | – | 605-612 |
| 80 | Autoimmune epilepsy | √ | √ | √ | √ | √ | 613-626 |

**Supplementary Table 3: Dermatological disorders mediated by B cells**

Involvement of B cells in the pathophysiology of autoimmune or immune-mediated diseases in dermatology through Ab-dependent and Ab-independent mechanisms, as supported by the presence of (auto)antibodies, (self)antigens, T cells, and/or cytokines. Reported benefit of rituximab is also noted for each indication. *Abbreviations*: PLC, pityriasis lichenoides chronica; PLEVA, pityriasis lichenoides et varioliformis acuta; FUMHD, febrile ulceronecrotic Mucha-Habermann disease.

| # | Indication | Antibodies | Antigens | T cells | Cytokines | Documented rituximab benefit | References |
|---|---|---|---|---|---|---|---|
| 81 | Pemphigus vulgaris | ✓ | ✓ | ✓ | ✓ | ✓ | 627-634 |
| 82 | Pemphigus foliaceus | ✓ | ✓ | ✓ | ✓ | ✓ | 629, 633, 635-638 |
| 83 | Mucous membrane pemphigoid | ✓ | ✓ | ✓ | ✓ | ✓ | 639-648 |
| 84 | Bullous pemphigoid | ✓ | ✓ | ✓ | ✓ | ✓ | 649-654 |
| 85 | Dermatitis herpetiformis | ✓ | ✓ | ✓ | ✓ | ✓ | 655-660 |
| 86 | Linear immunoglobulin A disease | ✓ | ✓ | ✓ | ✓ | ✓ | 661-667 |
| 87 | Epidermolysis bullosa acquisita | ✓ | ✓ | ✓ | ✓ | ✓ | 668-676 |
| 88 | Psoriasis | ✓ | ✓ | ✓ | ✓ | ✓ | 677-682 |
| 89 | Lichen planus | ✓ | ✓ | ✓ | ✓ | ✓ | 683-689 |
| 90 | Lichen sclerosus | ✓ | ✓ | ✓ | ✓ | – | 690-692 |
| 91 | Pityriasis rubra pilaris | ✓ | ✓ | ✓ | ✓ | – | 693-695 |
| 92 | Pityriasis lichenoides (PLC, PLEVA, FUMHD) | ✓ | ✓ | ✓ | ✓ | – | 696-700 |
| 93 | Atopic dermatitis | ✓ | ✓ | ✓ | ✓ | ✓ | 701-707 |
| 94 | Chronic autoimmune urticaria | ✓ | ✓ | ✓ | ✓ | ✓ | 708-713 |
| 95 | Autoimmune progesterone dermatitis | ✓ | ✓ | ✓ | ✓ | – | 714-720 |
| 96 | Weber-Christian disease | ✓ | ✓ | ✓ | ✓ | – | 721-724 |
| 97 | Pyoderma gangrenosum | – | – | ✓ | ✓ | ✓ | 725-728 |
| 98 | Hidradenitis suppurativa | ✓ | ✓ | ✓ | ✓ | – | 729-732 |
| 99 | Rosacea | ✓ | ✓ | ✓ | ✓ | – | 733-735 |
| 100 | Acne vulgaris | ✓ | ✓ | ✓ | ✓ | – | 735-740 |
| 101 | Vitiligo | ✓ | ✓ | ✓ | ✓ | ✓ | 741-748 |
| 102 | Alopecia areata | ✓ | ✓ | ✓ | ✓ | ✓ | 749-751 |
| 103 | Cutaneous lupus erythematosus | ✓ | ✓ | ✓ | ✓ | ✓ | 752-758 |

**Supplementary Table 4: Endocrine and metabolic disorders mediated by B cells**

Involvement of B cells in the pathophysiology of autoimmune or immune-mediated endocrine and metabolic disorders through Ab-dependent and Ab-independent mechanisms, as supported by the presence of (auto)antibodies, (self)antigens, T cells, and/or cytokines. Reported benefit of rituximab is also noted for each indication. *Abbreviations*: POEMS, polyneuropathy, organomegaly, endocrinopathy, monoclonal gammopathy, skin changes; IPEX, immune dysregulation, polyendocrinopathy, enteropathy, X-linked; ACTH, adrenocorticotropic hormone.

| # | Indication | Antibodies | Antigens | T cells | Cytokines | Documented rituximab benefit | References |
|---|---|---|---|---|---|---|---|
| 104 | Autoimmune polyglandular syndromes | ✓ | ✓ | ✓ | ✓ | ✓ | 759-762 |
| 105 | POEMS syndrome | ✓ | ✓ | ✓ | ✓ | – | 763-767 |
| 106 | IPEX syndrome | ✓ | ✓ | ✓ | ✓ | ✓ | 768-774 |
| 107 | Drug-induced autoimmunity | ✓ | ✓ | ✓ | ✓ | – | 775-779 |
| 108 | Addison disease | ✓ | ✓ | ✓ | ✓ | ✓ | 780-785 |
| 109 | Autoimmune adrenalitis | ✓ | ✓ | ✓ | ✓ | – | 786-792 |
| 110 | Isolated ACTH deficiency | ✓ | ✓ | – | – | – | 793-798 |
| 111 | Primary aldosteronism | ✓ | ✓ | ✓ | ✓ | – | 799-801 |
| 112 | Hashimoto’s disease | ✓ | ✓ | ✓ | ✓ | – | 802-807 |
| 113 | Graves’ disease | ✓ | ✓ | ✓ | ✓ | ✓ | 807-815 |
| 114 | Riedel thyroiditis | ✓ | ✓ | ✓ | ✓ | ✓ | 816-824 |
| 115 | Autoimmune hypoparathyroidism | ✓ | ✓ | ✓ | ✓ | – | 825-829 |
| 116 | Autoimmune parathyroiditis | ✓ | ✓ | ✓ | ✓ | – | 830-833 |
| 117 | Autoimmune hypophysitis | ✓ | ✓ | ✓ | ✓ | ✓ | 834-841 |
| 118 | Acromegaly | ✓ | ✓ | ✓ | ✓ | – | 842-848 |
| 119 | Diabetes mellitus type 1 | ✓ | ✓ | ✓ | ✓ | ✓ | 849-855 |
| 120 | Insulin autoimmune syndrome | ✓ | ✓ | ✓ | ✓ | ✓ | 856-863 |
| 121 | Type B insulin resistance syndrome | ✓ | ✓ | – | – | ✓ | 864-869 |
| 122 | Autoimmune hyperlipidemia | ✓ | ✓ | ✓ | ✓ | – | 870-881 |
| 123 | Light chain amyloidosis | ✓ | ✓ | ✓ | ✓ | – | 882-892 |

**Supplementary Table 5: Hematological and blood disorders mediated by B cells**

Involvement of B cells in the pathophysiology of autoimmune or immune-mediated diseases in hematology through Ab-dependent and Ab-independent mechanisms, as supported by the presence of (auto)antibodies, (self)antigens, T cells, and/or cytokines. Reported benefit of rituximab is also noted for each indication.

| # | Indication | Antibodies | Antigens | T cells | Cytokines | Documented rituximab benefit | References |
|---|---|---|---|---|---|---|---|
| 124 | Warm antibody hemolytic anemia | ✓ | ✓ | ✓ | ✓ | ✓ | 893-900 |
| 125 | Cold agglutinin disease | ✓ | ✓ | ✓ | ✓ | ✓ | 893, 894, 898-909 |
| 126 | Pure red blood cell aplasia | ✓ | ✓ | ✓ | ✓ | ✓ | 910-917 |
| 127 | Aplastic anemia | ✓ | ✓ | ✓ | ✓ | ✓ | 918-922 |
| 128 | Pernicious anemia | ✓ | ✓ | ✓ | ✓ | – | 923-925 |
| 129 | Evans syndrome | ✓ | ✓ | ✓ | ✓ | ✓ | 894, 897, 899, 926-930 |
| 130 | Autoimmune neutropenia & chronic idiopathic neutropenia | ✓ | ✓ | ✓ | ✓ | ✓ | 931-936 |
| 131 | Thrombotic thrombocytopenic purpura | ✓ | ✓ | ✓ | ✓ | ✓ | 937-944 |
| 132 | Immune thrombocytopenia | ✓ | ✓ | ✓ | ✓ | ✓ | 929, 930, 945-947 |
| 133 | Acquired hemophilia | ✓ | ✓ | ✓ | ✓ | ✓ | 948-953 |
| 134 | Antiphospholipid antibody syndrome | ✓ | ✓ | ✓ | ✓ | ✓ | 954-960 |
| 135 | Autoimmune lymphoproliferative syndrome | ✓ | ✓ | ✓ | ✓ | ✓ | 961-969 |
| 136 | Idiopathic multicentric Castleman disease | – | – | ✓ | ✓ | ✓ | 970-975 |
| 137 | Monoclonal gammopathy of undetermined significance | ✓ | ✓ | ✓ | ✓ | ✓ | 976-984 |
| 138 | Autoimmune myelofibrosis | ✓ | ✓ | ✓ | ✓ | – | 985-988 |

**Supplementary Table 6: Eye disorders mediated by B cells**

Involvement of B cells in the pathophysiology of autoimmune or immune-mediated diseases in ophthalmology through Ab-dependent and Ab-independent mechanisms, as supported by the presence of (auto)antibodies, (self)antigens, T cells, and/or cytokines. Reported benefit of rituximab is also noted for each indication.

| # | Indication | Antibodies | Antigens | T cells | Cytokines | Documented rituximab benefit | References |
|---|---|---|---|---|---|---|---|
| 139 | Autoimmune uveitis (iritis, vitritis, choroiditis, panuveitis) | ✓ | ✓ | ✓ | ✓ | ✓ | 989-1004 |
| 140 | Sympathetic ophthalmia | ✓ | ✓ | ✓ | ✓ | – | 1005-1010 |
| 141 | Vogt-Koyanagi-Harada disease | ✓ | ✓ | ✓ | ✓ | ✓ | 1006, 1011-1016 |
| 142 | Cogan syndrome | ✓ | ✓ | ✓ | ✓ | ✓ | 1017-1022 |
| 143 | Keratoconjunctivitis sicca | ✓ | ✓ | ✓ | ✓ | – | 1023-1031 |
| 144 | Peripheral ulcerative keratitis | ✓ | ✓ | ✓ | ✓ | – | 1032-1035 |
| 145 | Mooren's ulcer | ✓ | ✓ | ✓ | ✓ | ✓ | 1036-1042 |
| 146 | Scleritis & episcleritis | ✓ | ✓ | ✓ | ✓ | ✓ | 1043-1053 |
| 147 | Autoimmune ligneous conjunctivitis | ✓ | ✓ | ✓ | ✓ | – | 1054-1059 |
| 148 | Autoimmune retinopathy | ✓ | ✓ | ✓ | ✓ | ✓ | 1060-1068 |
| 149 | Posner-Schlossman syndrome | ✓ | ✓ | ✓ | ✓ | – | 1069-1071 |
| 150 | Orbital myositis | – | – | ✓ | ✓ | ✓ | 1072-1076 |
| 151 | Autoimmune dacryoadenitis | ✓ | ✓ | ✓ | ✓ | ✓ | 1077-1085 |

**Supplementary Table 7: Gastrointestinal disorders mediated by B cells**

Involvement of B cells in the pathophysiology of autoimmune or immune-mediated diseases in gastroenterology through Ab-dependent and Ab-independent mechanisms, as supported by the presence of (auto)antibodies, (self)antigens, T cells, and/or cytokines. Reported benefit of rituximab is also noted for each indication.

| # | Indication | Antibodies | Antigens | T cells | Cytokines | Documented rituximab benefit | References |
|---|---|---|---|---|---|---|---|
| 152 | Eosinophilic esophagitis | ✓ | ✓ | ✓ | ✓ | – | 1086-1091 |
| 153 | Autoimmune pancreatitis | ✓ | ✓ | ✓ | ✓ | ✓ | 1092-1099 |
| 154 | Autoimmune atrophic gastritis | ✓ | ✓ | ✓ | ✓ | – | 1100-1104 |
| 155 | Cronkhite-Canada syndrome | ✓ | ✓ | ✓ | ✓ | – | 1105-1111 |
| 156 | Autoimmune enteropathy | ✓ | ✓ | ✓ | ✓ | – | 1112-1121 |
| 157 | Celiac disease | ✓ | ✓ | ✓ | ✓ | – | 1122-1124 |
| 158 | Crohn’s disease | ✓ | ✓ | ✓ | ✓ | – | 1125-1131 |
| 159 | Ulcerative colitis | ✓ | ✓ | ✓ | ✓ | ✓ | 1126, 1128-1135 |
| 160 | Microscopic colitis | ✓ | ✓ | ✓ | ✓ | – | 1136-1143 |

**Supplementary Table 8: Kidney disorders mediated by B cells**

Involvement of B cells in the pathophysiology of autoimmune or immune-mediated diseases in nephrology through Ab-dependent and Ab-independent mechanisms, as supported by the presence of (auto) antibodies, (self)antigens, T cells, and/or cytokines. Reported benefit of rituximab is also noted for each indication.

| # | Indication | Antibodies | Antigens | T cells | Cytokines | Documented rituximab benefit | References |
|---|---|---|---|---|---|---|---|
| 161 | Lupus nephritis | ✓ | ✓ | ✓ | ✓ | ✓ | 1144-1149 |
| 162 | Idiopathic membranous nephropathy | ✓ | ✓ | ✓ | ✓ | ✓ | 1150-1159 |
| 163 | Immunoglobulin A nephropathy | ✓ | ✓ | ✓ | ✓ | ✓ | 1160-1167 |
| 164 | Membranoproliferative glomerulonephritis | ✓ | ✓ | ✓ | ✓ | ✓ | 1168-1177 |
| 165 | Minimal change disease | ✓ | ✓ | ✓ | ✓ | ✓ | 1178-1184 |
| 166 | Focal segmental glomerulosclerosis | ✓ | ✓ | ✓ | ✓ | ✓ | 1180-1182, 1185-1191 |
| 167 | Tubulointerstitial nephritis | ✓ | ✓ | ✓ | ✓ | – | 1192-1198 |

**Supplementary Table 9: Immunological and allergic disorders mediated by B cells**

Involvement of B cells in the pathophysiology of autoimmune or immune-mediated diseases in immunology through Ab-dependent and Ab-independent mechanisms, as supported by the presence of (auto)antibodies, (self)antigens, T cells, and/or cytokines. Reported benefit of rituximab is also noted for each indication.

| # | Indication | Antibodies | Antigens | T cells | Cytokines | Documented rituximab benefit | References |
|---|---|---|---|---|---|---|---|
| 168 | Primary antibody deficiency syndromes | ✓ | ✓ | ✓ | ✓ | ✓ | 1199-1207 |
| 169 | Common variable immunodeficiency | – | – | ✓ | ✓ | ✓ | 1207-1211 |
| 170 | Graft-versus-host disease | ✓ | ✓ | ✓ | ✓ | ✓ | 1212-1219 |
| 171 | Antibody-mediated rejection | ✓ | ✓ | ✓ | ✓ | ✓ | 1220-1230 |
| 172 | T cell-mediated rejection | ✓ | ✓ | ✓ | ✓ | ✓ | 1231-1237 |
| 173 | Acquired angioedema | ✓ | ✓ | ✓ | ✓ | ✓ | 1238-1244 |
| 174 | Drug-induced hypersensitivity syndrome | ✓ | ✓ | ✓ | ✓ | – | 1245-1255 |

**Supplementary Table 10: Cardiovascular disorders mediated by B cells**

Involvement of B cells in the pathophysiology of autoimmune or immune-mediated diseases in cardiology through Ab-dependent and Ab-independent mechanisms, as supported by the presence of (auto)antibodies, (self)antigens, T cells, and/or cytokines. Reported benefit of rituximab is also noted for each indication.

| # | Indication | Antibodies | Antigens | T cells | Cytokines | Documented rituximab benefit | References |
|---|---|---|---|---|---|---|---|
| 175 | Autoimmune myocarditis & dilated cardiomyopathy | ✓ | ✓ | ✓ | ✓ | ✓ | 1256-1269 |
| 176 | Post-cardiac injury syndrome (pericarditis) | ✓ | ✓ | ✓ | ✓ | ✓ | 1270-1281 |
| 177 | Autoimmune endocarditis | ✓ | ✓ | ✓ | ✓ | – | 1282-1290 |
| 178 | Atherosclerosis | ✓ | ✓ | ✓ | ✓ | – | 1291-1296 |
| 179 | Raynaud’s syndrome | ✓ | ✓ | ✓ | ✓ | ✓ | 1297-1304 |
| 180 | Chronic periaortitis | ✓ | ✓ | ✓ | ✓ | ✓ | 1305-1311 |
| 181 | Postural orthostatic tachycardia syndrome | ✓ | ✓ | ✓ | ✓ | – | 1312-1316 |

**Supplementary Table 11: Pulmonary disorders mediated by B cells**

Involvement of B cells in the pathophysiology of autoimmune or immune-mediated diseases in pulmonology through Ab-dependent and Ab-independent mechanisms, as supported by the presence of (auto)antibodies, (self)antigens, T cells, and/or cytokines. Reported benefit of rituximab is also noted for each indication. *Abbreviations*: IPF, idiopathic pulmonary fibrosis.

| # | Indication | Antibodies | Antigens | T cells | Cytokines | Documented rituximab benefit | References |
|---|---|---|---|---|---|---|---|
| 195 | Idiopathic pulmonary fibrosis | ✓ | ✓ | ✓ | ✓ | – | 1317-1322 |
| 196 | Interstitial lung diseases (non-IPF) | ✓ | ✓ | ✓ | ✓ | ✓ | 1323-1341 |
| 197 | Asthma | ✓ | ✓ | ✓ | ✓ | – | 1342-1346 |
| 198 | Goodpasture syndrome | ✓ | ✓ | ✓ | ✓ | ✓ | 1347-1355 |
| 199 | Sarcoidosis | ✓ | ✓ | ✓ | ✓ | ✓ | 1356-1364 |

**Supplementary Table 12: Hepatic and biliary disorders mediated by B cells**

Involvement of B cells in the pathophysiology of autoimmune or immune-mediated diseases in hepatology through Ab-dependent and Ab-independent mechanisms, as supported by the presence of (auto)antibodies, (self)antigens, T cells, and/or cytokines. Reported benefit of rituximab is also noted for each indication.

| # | Indication | Antibodies | Antigens | T cells | Cytokines | Documented rituximab benefit | References |
|---|---|---|---|---|---|---|---|
| 200 | Autoimmune hepatitis | ✓ | ✓ | ✓ | ✓ | ✓ | 1365-1371 |
| 201 | Primary biliary cholangitis | ✓ | ✓ | ✓ | ✓ | ✓ | 1372-1377 |
| 202 | Wilson disease | ✓ | ✓ | ✓ | ✓ | – | 1378-1381 |
| 203 | Primary sclerosing cholangitis | ✓ | ✓ | ✓ | ✓ | – | 1368, 1382-1386 |

**Supplementary Table 13: Psychiatric disorders mediated by B cells**

Involvement of B cells in the pathophysiology of autoimmune or immune-mediated diseases in psychiatry through Ab-dependent and Ab-independent mechanisms, as supported by the presence of (auto)antibodies, (self)antigens, T cells, and/or cytokines. Reported benefit of rituximab is also noted for each indication. *Abbreviations*: PANDAS, pediatric autoimmune neuropsychiatric disorders associated with streptococcal infections.

| # | Indication | Antibodies | Antigens | T cells | Cytokines | Documented rituximab benefit | References |
|---|---|---|---|---|---|---|---|
| 182 | Obsessive-compulsive disorder | ✓ | ✓ | ✓ | ✓ | – | 1387-1391 |
| 183 | PANDAS | ✓ | ✓ | ✓ | ✓ | – | 1392-1397 |
| 184 | Autoimmune psychosis | ✓ | ✓ | ✓ | ✓ | – | 1398-1404 |
| 185 | Schizophrenia | ✓ | ✓ | ✓ | ✓ | – | 1405-1415 |
| 186 | Bipolar disorder | ✓ | ✓ | ✓ | ✓ | – | 1416-1425 |
| 187 | Borderline personality disorder | ✓ | ✓ | – | ✓ | – | 1424, 1426-1428 |
| 188 | Autism spectrum disorder | ✓ | ✓ | ✓ | ✓ | – | 1429-1440 |

**Supplementary Table 14: Gynecological disorders mediated by B cells**

Involvement of B cells in the pathophysiology of autoimmune or immune-mediated diseases in gynecology through Ab-dependent and Ab-independent mechanisms, as supported by the presence of (auto)antibodies, (self)antigens, T cells, and/or cytokines. Reported benefit of rituximab is also noted for each indication.

| # | Indication | Antibodies | Antigens | T cells | Cytokines | Documented rituximab benefit | References |
|---|---|---|---|---|---|---|---|
| 189 | Primary ovarian insufficiency | ✓ | ✓ | ✓ | ✓ | – | 1441-1447 |
| 190 | Autoimmune oophoritis | ✓ | ✓ | ✓ | ✓ | – | 1448-1453 |
| 191 | Polycystic ovary syndrome | ✓ | ✓ | ✓ | ✓ | – | 1454-1459 |
| 192 | Pemphigoid gestationis | ✓ | ✓ | ✓ | ✓ | ✓ | 1460-1470 |
| 193 | Autoimmune congenital heart block | ✓ | ✓ | ✓ | ✓ | – | 1471-1477 |
| 194 | Endometriosis | ✓ | ✓ | ✓ | ✓ | – | 1478-1485 |

**Supplementary Table 15: Ear, nose, and throat disorders mediated by B cells**

Involvement of B cells in the pathophysiology of autoimmune or immune-mediated diseases in otolaryngology through Ab-dependent and Ab-independent mechanisms, as supported by the presence of (auto)antibodies, (self)antigens, T cells, and/or cytokines. Reported benefit of rituximab is also noted for each indication.

| # | Indication | Antibodies | Antigens | T cells | Cytokines | Documented rituximab benefit | References |
|---|---|---|---|---|---|---|---|
| 204 | Autoimmune vertigo | ✓ | ✓ | ✓ | ✓ | – | 1486,1487 |
| 205 | Idiopathic bilateral vestibulopathy | ✓ | ✓ | – | – | – | 1488,1489 |
| 206 | Autoimmune sensorineural hearing loss | ✓ | ✓ | ✓ | ✓ | ✓ | 1490-1493 |
| 207 | Ménière’s disease | ✓ | ✓ | – | – | – | 1494-1496 |

**Supplementary Table 16: Genitourinary disorders mediated by B cells**

Involvement of B cells in the pathophysiology of autoimmune or immune-mediated diseases in urology through Ab-dependent and Ab-independent mechanisms, as supported by the presence of (auto)antibodies, (self)antigens, T cells, and/or cytokines. Reported benefit of rituximab is also noted for each indication.

| # | Indication | Antibodies | Antigens | T cells | Cytokines | Documented rituximab benefit | References |
|---|---|---|---|---|---|---|---|
| 208 | Chronic prostatitis (chronic pelvic pain syndrome) | ✓ | ✓ | ✓ | ✓ | – | 1497-1503 |
| 209 | Interstitial cystitis | ✓ | ✓ | ✓ | ✓ | – | 1504-1507 |
| 210 | Autoimmune orchitis | ✓ | ✓ | ✓ | ✓ | – | 1508-1513 |
| 211 | Autoimmune male infertility | ✓ | ✓ | ✓ | ✓ | – | 1514-1523 |

**Supplementary Table 17: Infectious diseases mediated by B cells**

Involvement of B cells in the pathophysiology of autoimmune or immune-mediated diseases in infectiology through Ab-dependent and Ab-independent mechanisms, as supported by the presence of (auto)antibodies, (self) antigens, T cells, and/or cytokines. Reported benefit of rituximab is also noted for each indication.

| # | Indication | Antibodies | Antigens | T cells | Cytokines | Documented rituximab benefit | References |
|---|---|---|---|---|---|---|---|
| 212 | Chagas disease | ✓ | ✓ | ✓ | ✓ | – | 1524-1526 |
| 213 | Lyme disease | ✓ | ✓ | ✓ | ✓ | – | 1527-1530 |
